# Whole genome sequencing of a family with autosomal dominant features within the oculoauriculovertebral spectrum

**DOI:** 10.1101/2024.02.07.24301824

**Authors:** AL Petrin, LA Machado-Paula, A Hinkle, L Hovey, W Awotoye, M Chimenti, B Darbro, LA Ribeiro-Bicudo, SM Dabdoub, T Peter, P Breheny, J Murray, E Van Otterloo, S Rengasamy Venugopalan, LM Moreno-Uribe

**Author notes:** Corresponding author: Aline L Petrin, Department of Orthodontics, Iowa Institute of Oral Health Research, College of Dentistry and Dental Clinics, University of Iowa, Iowa City, IA, USA.

## Abstract

**Background:** Oculoauriculovertebral Spectrum (OAVS) encompasses abnormalities on derivatives from the first and second pharyngeal arches including macrostomia, hemifacial microsomia, micrognathia, preauricular tags, ocular and vertebral anomalies. We present genetic findings on a three-generation family affected with macrostomia, preauricular tags and uni- or bilateral ptosis following an autosomal dominant pattern.

**Methods:** We generated whole genome sequencing data for the proband, affected parent and unaffected paternal grandparent followed by Sanger sequencing on 23 family members for the top 10 candidate genes: *KCND2, PDGFRA, CASP9, NCOA3, WNT10A, SIX1, MTF1, KDR/VEGFR2, LRRK1,* and *TRIM2* We performed parent and sibling-based transmission disequilibrium tests and burden analysis via a penalized linear mixed model, for segregation and mutation burden respectively. Next, via bioinformatic tools we predicted protein function, mutation pathogenicity and pathway enrichment to investigate the biological relevance of mutations identified.

**Results:** Rare missense mutations in *SIX1, KDR/VEGFR2,* and *PDGFRA* showed the best segregation with the OAV phenotypes in this family. When considering any of the 3 OAVS phenotypes as an outcome, *SIX1* had the strongest associations in parent-TDTs and sib-TDTs (p=0.025, p=0.052) (unadjusted p-values). Burden analysis identified *SIX1* (RC=0.87) and *PDGFRA* (RC=0.98) strongly associated with OAVS severity. Using phenotype-specific outcomes, sib-TDTs identified *SIX1* with uni- or bilateral ptosis (p=0.049) and ear tags (p=0.01), and *PDGFRA* and *KDR/VEGFR2* with ear tags (both p<0.01).

**Conclusion:** *SIX1*, *PDGFRA*, and *KDR/VEGFR2* are strongly associated to OAVS phenotypes. *SIX1* has been previously associated with OAVS ear malformations and is co-expressed with *EYA1* during ear development. Efforts to strengthen the genotype-phenotype co-relation underlying the OAVS are key to discover etiology, family counseling and prevention.

## Introduction

Oculoauriculovertebral Spectrum (OAVS) encompasses a wide variety of anomalies on structures derived from the first and second pharyngeal arches, including macrostomia, hemifacial microsomia, micrognathia, preauricular tags, ocular and vertebral anomalies ^1^. Although the specific etiology of OAVS anomalies is still elusive the literature describes 3 pathogenic and possibly interacting models that may in part explain craniofacial anomalies, yet not the postcranial findings also described including lung, heart, kidney, skeletal and gastrointestinal anomalies^2^. The pathological models include vascular abnormalities and hemorrhage, abnormal development of cranial neuro crest cells (CNCCs) affecting their migration, proliferation or differentiation and damage to Meckel’s cartilage ^3^. Healthy dynamics of CNCCs and tissue derivatives within the pharyngeal arches are key during the fourth and eight weeks of embryonic development, a critical stage in human facial formation. During the 5^th^ week, the growing stomodeum is surrounded by multiple, pertinent areas of differentiating cells like the mandibular and maxillary processes, and median and lateral nasal processes that grow, fuse, and differentiate giving rise to the maxillomandibular complex^4^.

One of the common findings within the OAVS is macrostomia (Tessier cleft type 7) which accounts for 0.3-1% of facial clefts and is found in about 16-36% of cases with other OAV features ^5^. Macrostomia occurs due to the failure in fusion of the maxillary and mandibular processes during week 6^th^ of development. The orotragal line, an embryological line extending from the lateral commissure of the stomodeum to the tragus of the ear, is meant to dissipate once the maxillary and mandibular processes fuse. If fusion fails, then the line continues to develop into the abnormal enlargement of the labial commissure ^4^. Anatomically, the abnormal contour of the mouth commissure varies from a mild lateral displacement to a complete transverse cleft to the ear. The orbicularis oris muscle abnormally terminates at this commissure, causing less distance between its location and the tragus ^4, 6^. Macrostomia has an incidence of 1 in 60,000 to 1 in 300,000 live births, with more common occurrence in males ^7, 8^. While it may occur unilaterally or bilaterally, unilateral cases are more frequent with the right side more often affected than the left side ^9^.

Although less frequently, the presence of uni- or bilateral ptosis or drooping of the upper eyelid has also been reported in cases within the OAVS ^10, 11^. The phenotype can be due defects in the innervation of levator palpebrase superioris muscle, which starts to develop during the fifth week of gestation. It has also been suggested that the OAVS may include defects of the otic placode ^1^. The phenotypic complexity of the OAVS, with the involvement of multisystem anomalies suggests that more than one etiological variant is at play.

While the embryological development of the OAVS abnormalities is well understood, the etiology of the defects is not. Environmental factors such as exposure to toxic substances during pregnancy ^12, 13^, maternal diabetes, and hemorrhage ^14^ have been associated with OAVS, as well as epigenetic dysregulations ^15^.

As for the genetic component of OAVS complex etiology, with the advances of next generation sequencing, several genes have been associated however, their pathophysiology remains underexplored. The first gene to be associated with OAVS was *MYT1* ^16–19^; after which several others studies reported additional genes, *SF3B2* ^20^, *ZYG11B* ^21^, *EYA3* ^22, 23^, *VWA1* ^24^, *ZIC3* ^25^, *AMIGO2* ^26^, *OTX2*, *YPEL1* ^27^, PTCH2 ^28^.

Structural variants in 4p16, 14q22.3, and 22q11.2 among other loci have also been described ^29–38^. Although most causal mutations and structural variants in these genes/loci occur *de novo,* familial inheritance has also been reported, mostly with autosomal dominant inheritance ^20, 23, 39–41^.

In 2006, researchers Richieri-Costa et. al. investigated a three-generational family with multiple members experiencing macrostomia, preauricular tags, uni- or bilateral ptosis, and external ophthalmoplegia ^1^. We report an updated pedigree of that family and molecular data using DNA samples from some affected and unaffected family members. First, we generated whole genome sequencing (WGS) data for the affected proband, the affected father, and the unaffected paternal grandmother. Then we followed with Sanger sequencing to investigate segregation of the best candidate genes mutations in the additional available family members.

The objective of this study was to determine the gene(s) that harbor the pathogenic variant that led to the OAVS of abnormalities in the family. These findings support the role of a complex gene network behind craniofacial development and provide additional information for understanding and strengthening of the genotype-phenotype correlations underlying the phenotypic variability present in cases with OAVS.

## Methods

### Patients

The detailed clinical description of the family was previously published by Richieri-Costa et al (2006) ^1^. Briefly, the female proposita presented at birth with bilateral macrostomia, preauricular sinus, and preauricular tags. The parent presented with normal mouth contour and preauricular tags. Other members of the family present one or more of the following: macrostomia, preauricular skin tags, and/or uni- or bilateral ptosis*. As previously reported by Richieri-Costa et al (2006) ^1^, the examined family members had no apparent neurological symptoms, abnormal ocular movement, bulbar signs, limb anomalies, or muscle weakness in the extremities at the time of examination^1^. We obtained DNA samples from 8 affected and 15 unaffected family members across the three generations. The study was approved by the respective institutional review boards and all participants signed informed consents prior collection of clinical data and biological samples.

### Whole Genome Sequencing and Variant Calling

We generated whole-genome sequencing (WGS) data (30X coverage) for the proband, affected parent and unaffected paternal grandparent and analyzed the entire genome of the three individuals for pathogenic protein-altering genetic variants that were shared between the proband and the parent but not carried by the unaffected grandparent. Pathogenic variants that segregated within the affected individuals (proband and parent) but not in control (grandparent) were further investigated within the expanded family.

*Genomic Analysis workflow:* The genomes of the three selected individuals were sequenced at an average coverage depth of 30x. The sequence data were mapped and aligned to the human genome assembly GRCh38 (Hg38). Following the alignment, alternate alleles at each genomic locus were called using the highly performing Dynamic Read Analysis for GENomics (DRAGEN) pipeline ^42, 43^. These alternate alleles included single nucleotide variants (SNVs), insertions and deletions (InDels <1Kb), and copy number variation (deletions and duplications >1Kb). For this publication, we only focused on analyses and segregation of SNVs. Structural variant analyses of this pedigree will be the subject of a future publication.

As a quality control measures, we selected variants with a genotype quality (GQ) of at least 20 and a read depth (RD) of at least 10. Next, we prioritized rare protein-altering genetic variants. These rare variants were selected based on ^†^minor allele frequency (MAF) less than 1% (0.01) reported on the genomic population database, gnomAD (https://gnomad.broadinstitute.org/). This pipeline is based on the hypothesis that this familial craniofacial condition is caused by rare protein-altering genetic variant(s). Next, we used the Mouse Genomics Informatics (MGI) Database (https://www.informatics.jax.org) to investigate the association between the genes harboring identified pathogenic variants and craniofacial phenotypes observed in mice prioritizing those similar to the reported traits within the family. We included all variants with gnomAD constraint scores less than 0.9 and z scores greater than 3.5 for LOF and missense, respectively, and also included genes with significant craniofacial phenotype in the MGI database.

Next, we used *in silico* tools including Sorting Intolerant From Tolerant, SIFT (http://sift.jcvi.org/), Polymorphism Phenotyping, PolyPhen2^44^ (http://genetics.bwh.harvard.edu/pph2/), Combined Annotation Dependent Depletion – CADD score^45^ - (https://cadd.gs.washington.edu/) and Varsome^46^ – (https://varsome.com/) to predict the pathogenicity of the selected protein-altering variants. In addition, we used the web-based machine-learning algorithm, DOMINO (https://domino.iob.ch/) to assess the probability that the genes harboring these variants are likely to cause dominant changes, thus matching the inheritance pattern of the craniofacial condition in the family.

### Sanger Sequencing

We designed primers for mutations located in the top 10 candidate genes (*KCND2, PDGFRA, CASP9, NCOA3, WNT10A, SIX1, MTF1, KDR/VEGFR2, LRRK1,* and *TRIM2)* that resulted from the pipeline described above and sequenced DNA from 23 available family members (affected and unaffected individuals) to investigate the segregation of each mutation. Primer sequences and PCR conditions are available upon request. PCR products were verified in agarose gel and submitted for Sanger sequencing at the University of Iowa DNA Core facility.

### Statistical Analysis

After confirming the presence or absence of the mutations in the 23 individuals, we performed three analyses: (1) parent and sibling-based transmission disequilibrium tests (TDTs) to examine associations with any OAVS anomaly, (2) a burden analysis to explore the effects of the candidate gene mutations on OAVS phenotype severity, and (3) parent and sibling-based TDTs to detect associations with specific OAVS phenotypes.

For our first analysis, we classified each individual as being ’affected’ or ’unaffected’ based on the presence/absence of OAVS abnormalities. Individuals with any one of or any combination of macrostomia, uni- or bilateral ptosis, or ear tags were designated as ’affected.’ One of the 23 individuals had an indeterminate phenotype and was believed to be a non-penetrant obligated carrier based on the genotype and family data. The present analysis designated this individual of indeterminate phenotype as ’unaffected,’ keeping consistent with previous publications based on this data set. Two statistical tests were performed: the parent-TDT ^47^ and the sib-TDT ^48^, both of which are variations of the well-known transmission disequilibrium test (TDT). The parent-TDT combines the traditional TDT with a parental discordance test, thereby treating each nuclear family parental couple as a matched pair in addition to analyzing parent-child trios. The sib-TDT combines the traditional TDT with a sibling-based method, so that unaffected siblings can serve as controls for their affected siblings when genotype data from parents is unavailable. A total of 18 tests of association were implemented using PLINK software (version 1.9).

In the second analysis (burden test), we defined a numeric outcome indicating the number of OAVS anomalies present on an individual. This value ranged from 0 to 3, with 0 representing individuals with no OAVS anomalies and 3 representing individuals with a maximum number of 3 anomalies and therefore greater phenotypic severity. With this outcome, we used a penalized linear mixed modelling (PLMM) approach ^49^ to identify the genes with the strongest evidence of association with the severity of OAVS phenotypes. This PLMM approach incorporated all the individuals in the data, accounting for the relatedness among family members using the average percentage of DNA shared between relatives (23andMe, 2022). PLMM does not perform association tests for each gene; instead, this method treats the candidate gene mutations as one system and then chooses the members of the system which have the most evidence of association with the severity of the phenotype. PLMM was implemented in R (version 4.3) using the package plmmr (version 1.1) ^50^.

For the third analysis, we used parent and sibling based TDTs to generate hypotheses about associations between individual candidate gene mutations and specific OAVS phenotypes. This involved a total of 27 tests, one for each combination of gene and OAVS phenotype.

Since the goals across all three of our analyses were exploratory, no p-values were adjusted for multiple comparisons.

### Bioinformatic Analysis

The InterPro database was used to investigate the specific candidate proteins, their domains, and functional sites ^51^. For prediction of the potential pathogenic potential of 3D protein structure alterations due to mutations, we used AlphaMissense ^52^. Finally, we used MobiDB to investigate protein disorder and mobility annotations ^53^. Active subnetwork pathway enrichment analysis of genes determined to be significantly associated (see previous section) was carried out using the R package pathfindR v2.2.0^54^. Briefly, the list of genes and p-values was loaded as input, genes were filtered through the Biogrid protein interaction network ^55, 56^ and the resulting set of interactions (with at most one indirect interaction) were used for pathway enrichment analysis through the GO ^57^ and Reactome ^58^ databases.

## Results

### Whole genome Sequencing

To identify novel or rare protein-altering variants, we extracted high confidence LoF (stop-gained, frameshift, and splice acceptor/donor) and missense variants with minor allele frequency ≤1% when compared with the human global and ethnic -specific database (gnomAD) ^59^. Following our data analysis pipeline, we identified 14 loss of function (LOF) and 222 missense mutations

(Supplemental Table 1) that were present in both the parent and proband but not present in the unaffected grandparent as illustrated in Figure 2.

**Figure 2:**
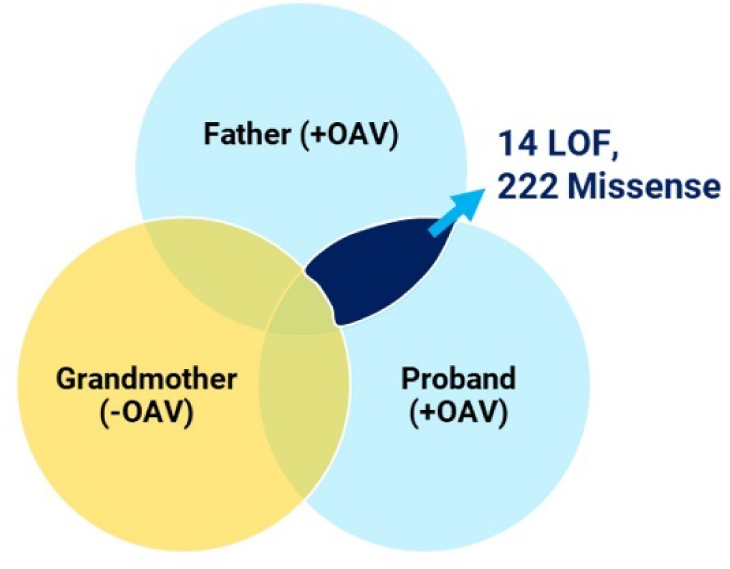
14 LOF and 222 Missense genes were shared solely between the father (+OAV) and proband (+OAV)

From those, we kept variants with gnomAD constraint scores less than 0.9 and z score greater than 3.5 for LOF and missense (3 genes in Table 1), respectively; and included genes with significant craniofacial phenotype in the MGI database (13 genes in Table 2).

**Table 1:** Resulting genes based on gnomAD constraint metrics.

| Gene of Interest | Chr. Pos (hg38) | Effect | Missense Z | pLoF o/e |
| --- | --- | --- | --- | --- |
| *KCND2* | 7:120733051 | LoF | 4.65 | 0.43 |
| *UNC80* | 2:209819105 | Missense | 4.99 | 0.48 |
| *TRIM2* | 4:153257397 | Missense | 3.94 | 0.4 |

**Table 2:** Resulting genes associated with human and/or mouse craniofacial phenotype based on the Mouse Genomics Informatics Database.

| Gene | Chr. Pos. (hg38) | Mutation Effect | Human Phenotypes | Mouse Phenotypes |
| --- | --- | --- | --- | --- |
| *KIAA0586* | 14:58470656 | Missense | Cleft palate, Depressed Nasal Bridge | Abnormal lateral/medial nasal prominence, narrow head |
| *NCOA3* | 20:47651093 | Missense | NA | Abnormal tooth morphology, lower ear position |
| *CASP9* | 1:15518262 | Missense | Hyposmia | Abnormal olfactory epithelium |
| *PDGFRA* | 4:54288865 | Missense | NA | Cleft palate, impaired ossification of basisphenoid bone, decreased cranium height, abnormal frontal/parietal/nasal bone morphology, absent zygomatic bone, ectopic cranial bone, abnormal frontonasal prominence, abnormal nasal capsule, short snout, midline facial cleft |
| *WNT10A* | 2:218890256 | Missense | Shopf-Shulz-Passarge Syndrome, tooth agenesis | Abnormal alveolar process/incisor/molar morphology, dental pulp stones, taurodontia, abnormal dentin morphology, abnormal tooth resorption, abnormal tongue morphology |
| *FER* | 5:108954852 | Missense | NA | Abnormal snout morphology |
| *KDR/VEGFR2* | 4:55104697 | Missense | NA | Abnormal mandibular prominence morphology, absent third/fourth pharyngeal arch, abnormal second pharyngeal arch |
| *SIX1* | 14:60646457 | Missense | Preauricular Pit | Abnormal craniofacial morph, abnormal craniofacial bone morph, abnormal styloid process/Meckel's cartilage/frontal bone morph, short temporal bone squamous part, abnormal hyoid bone greater horn morphology, short mandible, short maxilla, micrognathia, abnormal nasal bone morph, absent turbinates, short zygomatic bone, abnormal incus/malleus/stapes morph, absent stapes, abnormal face development, abnormal midface morph, abnormal genioglossus muscle, dec tongue size, abnormal nose morph, abnormal external nares morph, abnormal nasal cavity morph, absent vomeronasal organ, abnormal/absent olfactory epithelium |
| *KDF1* | 1:26951399 | Missense | Short philtrum (H), natal tooth (H), concave nasal ridge (H) | NA |
| *MTF1* | 1:37857470 | Missense | NA | Palatal shelf fusion with tongue, cleft palate, abnormal tongue epithelium morphology, oral atresia, abnormal nose morphology, short snout |
| *NPHP3* | 3:132708187 | Missense | Meckel syndrome, renal hepatic pancreatic dysplasia, large fontanelles, dandy walker malformation, potter facies, preauricular<br>pit, high forehead | Premature tooth eruption, malocclusion |
| *DUOX2* | 15:45105795 | Missense | NA | Abnormal cranium morphology, abnormal zygomatic bone morphology, abnormal snout morphology |
| *LRRK1* | 15:101010802 | Missense | NA | Abnormal tooth morphology, absent teeth, abnormal mandible morphology, abnormal maxilla morph, abnormal zygomatic morph, abnormal facial morph, abnormal snout morph |

Lastly, based on the autosomal dominant inheritance in this family, we used the Domino Database to assess inheritance patterns of the 16 genes that resulted from the filtering steps above. Any genes that had extremely low odds of harboring an autosomal dominant mutation were excluded from candidacy. Only “Likely dominant” to “Very likely dominant” were considered as candidates to harbor etiological variants, which narrowed our list of candidates down to 10 genes (Table 3).

**Table 3:** Genes with very likely or likely dominant inherited based on Domino Database.

| Gene of Interest | Mutation | LDA Score | P(AD) | Class |
| --- | --- | --- | --- | --- |
| *SIX1* | D227E | 3.698 | 1 | Very likely dominant |
| *KCND2* | R422* | 3.172 | 1 | Very likely dominant |
| *PDGFRA* | R914Q | 2.857 | 0.999 | Very likely dominant |
| *MTF1* | D63E | 2.47 | 0.997 | Very likely dominant |
| *KDR/VEGFR2* | Q645E | 2.336 | 0.996 | Very likely dominant |
| *NCOA3* | Q1276del | 2.235 | 0.994 | Very likely dominant |
| *LRRK1* | L416M | 1.104 | 0.833 | Very likely dominant |
| *CASP9* | L89P | 0.836 | 0.718 | Likely dominant |
| *WNT10A* | D217N | 0.655 | 0.624 | Likely dominant |
| *TRIM2* | 12941G>A | 0.642 | 0.617 | Likely dominant |

P(AD) - Probabilities of being dominant

### Segregation Analysis of top 10 candidate genes

Mutations found in the 10 candidate genes that resulted in a likely dominant or very likely dominant were then tested for segregation in additional family members We designed primers specific for each mutation and used Sanger sequencing to confirm presence or absence of the mutations in all available family members. We used TDT tests to examine associations between each gene and the binarized phenotype ‘affected/unaffected’, where affected was defined as having any number of OAVS anomalies. Moreover, we used a penalized regression method to analyze the genes together as a system, where the outcome was the burden of OAVS anomalies. The burden outcome was defined as a number on a scale, ranging from 0 (no OAVS phenotypes) to 3 (the proposita, who has all 3 OAVS phenotypes). Of the 23 family members represented in this study, 6 presented uni- or bilateral ptosis, 6 presented ear tags, and 2 presented macrostomia and a total of 5 individuals presented multiple OAVS phenotypes. It is important to mention that there were an additional 5 individuals in the original pedigree who were described as presenting an abnormal mouth phenotype described as an atypical angle of the mouth with the lower lip vermillion shorter than the upper lip and a broad and irregular commissure^1^. For the purposes of these analyses, only the two individuals with the most severe mouth phenotypes (III-1 (macrostomia) and III-5 (wide mouth) were considered as affected with macrostomia. The results are shown in Table 4.

**Table 4:** Results from parent-TDT, sibling-TDT tests and burden analysis.

| Table 4: Results from parent-TDT, sibling-TDT tests and burden analysis |  |  |  |  |  |  |  |  |
| --- | --- | --- | --- | --- | --- | --- | --- | --- |
| Chr | Gene | Parent TDT |  |  | Sibling TDT |  |  | Burden analysis |
|  |  | transmitted counts | untransmitted counts | P_par | observed minor alleles | expected minor alleles | P_sib | plmm_coefficient |
| 1 | *CASP9* | 1 | 0 | 0.16 | 5 | 4.12 | 0.37 | 0 |
| 1 | *MTF1* | 1 | 0 | 0.08 | 6 | 5.26 | 0.38 | 0 |
| 2 | *WNT10A* | 1 | 0 | 0.16 | 5 | 4.12 | 0.37 | 0 |
| 4 | *PDGFRA* | 1 | 1 | 0.18 | 7 | 4.62 | 0.07 | 0.98 |
| 4 | *KDR* | 1 | 1 | 0.18 | 7 | 4.62 | 0.07 | 0 |
| 4 | *TRIM2* | 2 | 1 | 0.18 | 6 | 5.6 | 0.76 | 0 |
| 7 | *KCND2* | 1 | 0 | 0.16 | 5 | 4.12 | 0.37 | 0 |
| 14 | *SIX1* | 2 | 0 | 0.03 | 7 | 4.52 | 0.05 | 0.87 |
| 15 | *LRRK1* | 1 | 1 | 0.32 | 5 | 4.05 | 0.47 | 0.08 |

Note: p-values are not adjusted for multiple comparisons; transmitted\_counts: Transmitted minor allele count; untransmitted\_counts: Untransmitted allele count; P\_par: Combined test asymptotic p-value; observed\_minor\_alleles: Number of observed minor alleles; expected\_minor\_alleles: Number of expected minor alleles; P\_sib: Asymptotic p-value; plmm\_coefficient: Regression coefficient (i.e., beta 'hat')

To examine any associations between each OAV phenotype and each gene, we performed an exploratory analysis to discern whether some genes are more closely associated with a specific OAV phenotype. The sib-TDT with each of the nine genes in combination with each of the 3 phenotypes (ear tags, ptosis, and macrostomia) (Table 5) indicate that mutations in genes *PDGFRA* and *KDR/VEGFR2* were associated with the ear tags phenotype. We noted that *PDGFRA* and *KDR/VEGFR2* are in high linkage disequilibrium and thus, all individuals with a mutation in *PDGFRA* also have a mutation in *KDR/VEGFR2*.

**Table 5:** Results from phenotype specific TDTs.

| Table 5: Results from phenotype specific TDTs |  |  |  |  |  |
| --- | --- | --- | --- | --- | --- |
| CHR | Gene | A1 | A2 | P | Phenotype |
| 1 | *CASP9* | G | A | 0.96 | Uni- or bilateral ptosis |
| 1 | *MTF1* | A | C | 0.89 | Uni- or bilateral ptosis |
| 2 | *WNT10A* | A | G | 0.96 | Uni- or bilateral ptosis |
| 4 | *PDGFRA* | A | G | 0.2 | Uni- or bilateral ptosis |
| 4 | *KDR/VEGFR2* | G | C | 0.2 | Uni- or bilateral ptosis |
| 4 | *TRIM2* | A | G | 0.5 | Uni- or bilateral ptosis |
| 7 | *KCND2* | T | C | 0.96 | Uni- or bilateral ptosis |
| **14** | ***SIX1*** | **T** | **C** | **0.05** | **Uni- or bilateral ptosis** |
| 15 | *LRRK1* | A | C | 0.36 | Uni- or bilateral ptosis |
| 1 | *CASP9* | G | A | 0.58 | Ear Tags |
| 1 | *MTF1* | A | C | 0.63 | Ear Tags |
| 2 | *WNT10A* | A | G | 0.58 | Ear Tags |
| **4** | ***PDGFRA*** | **A** | **G** | **<0.01** | **Ear Tags** |
| **4** | ***KDR/VEGFR2*** | **G** | **C** | **<0.01** | **Ear Tags** |
| 4 | *TRIM2* | A | G | 0.81 | Ear Tags |
| 7 | *KCND2* | T | C | 0.58 | Ear Tags |
| **14** | ***SIX1*** | **T** | **C** | **0.01** | **Ear Tags** |
| 15 | *LRRK1* | A | C | 0.9 | Ear Tags |
| 1 | *CASP9* | G | A | 0.16 | Macrostomia |
| 1 | *MTF1* | A | C | 0.16 | Macrostomia |
| 2 | *WNT10A* | A | G | 0.16 | Macrostomia |
| 4 | *PDGFRA* | A | G | 0.16 | Macrostomia |
| 4 | *KDR/VEGFR2* | G | C | 0.16 | Macrostomia |
| 4 | *TRIM2* | A | G | 0.48 | Macrostomia |
| 7 | *KCND2* | T | C | 0.16 | Macrostomia |
| 14 | *SIX1* | T | C | 0.48 | Macrostomia |
| 15 | *LRRK1* | A | C | 0.16 | Macrostomia |

Bold indicates significant association with p-values<0.05

The data also show that there is nominal association (P<0.05) between mutations in the *SIX1* gene and the ear tag phenotype. In the tests examining the uni- or bilateral ptosis phenotype, the mutation in *SIX1* had the strongest association compared to other candidate gene mutations. No genetic mutations showed an association with macrostomia at this time. Future analyses including a broader definition of the macrostomia phenotype will be considered to assess the impact of these mutations on the macrostomia spectrum.

### Bioinformatic

Following the statistical analysis identifying *SIX1, PDGFRA,* and *KDR/VEGFR2/ VEGFR2* as significantly associated mutations; we investigated the potential functional implications of these mutations.

*PDGFRA* (platelet derived growth factor receptor alpha) is a cell surface tyrosine kinase receptor. As listed in the InterPro classification, while the mutation site identified in this study is not part of the active site (p.814-p.826), it does come in at the end of the overall kinase domain (p.593-p.954). To further dig into this mutation, we turned to AlphaMissense ^52^ which trained a two-stage neural network to incorporate predicted 3D structure (similar to AlphaFold) and abundance of known human proteins and calibrated against the ClinVar database of known relationships between protein variants and human health conditions to generate a predicted probability of the input variant being pathogenic. For the p.R914Q mutation, AlphaMissense classifies it as having an 80% probability of pathogenicity (along with a mean 93% probability of all mutations at that site).

Turning to *SIX1*, a transcription factor involved in embryonic development and often co-expressed with *EYA1*, InterPro indicates that p.217 does not occur within the primary homeodomain. However, MobiDB, a *database* of protein disorder and mobility, predicts that p.227 exists within a Linear Interacting Peptide (LIP) region, as well as a 99bp Intrinsically Disordered Region (IDR). Such proteins do not attain “stable three-dimensional structure under physiological conditions” and can display a high degree of conformational flexibility that endows proteins or protein regions with functional promiscuity ^60^. When we look at the exchange of Aspartic Acid for Glutamic acid in the p.D227E mutation, we see a move to a higher molecular weight residue with a larger van der Waals volume. Combined with the fact that LIP regions are responsible for mediating protein-protein interactions and the mutation further being within an IDR, indicates a role for this mutation to interfere with protein flexibility and potentially its regulatory function.

Finally, *KDR/VEGFR2/* (vascular endothelial growth factor receptor 2) encodes a kinase insert domain receptor of *VEGF*(A/C/D) that is essential for vascular development and hematopoiesis [Uniprot P35968]. InterPro indicates that p.645 occurs within a protein kinase domain (similar to *PDGFRA*) and Q645E is a known missense mutation listed in COSMIC (Catalogue of Somatic Mutations in Cancer) [COSV55760314] that affects the VEGF signaling, angiogenesis, and focal adhesion pathways.

Moving beyond the implications of individual genes/proteins, we turned to pathway overrepresentation analysis through the R package pathfindR. This method was chosen specifically because pathfindR, prior to performing pathway analysis, filters the input genes through databases of protein-protein interaction networks (PIN) to ensure validated interactions with at most one intervening protein. Using the Biogrid PIN we identified connections between *KDR/VEGFR2* and *PDGFRA* with *CAV1* as an intermediary implicated in GO pathways for peptidyl-tyrosine phosphorylation (GO:0018108), positive regulation of phosphatidylinositol 3-kinase signaling (GO:0014068), and transmembrane receptor protein tyrosine kinase activity (GO:0004714). Furthermore, *SIX1* and *PDGFRA* were connected by FZR1 in the embryonic cranial skeleton morphogenesis (GO:0048701) pathway. Finally, *KDR/VEGFR2* and *PDGFRA* were connected in the Reactome database within the signaling by *PDGFRA* in disease pathway (R-HSA-9671555), further validating the AlphaMissense results.

## Discussion

OAVS is the second most common craniofacial birth defect. Rehabilitation of the wide spectrum of anomalies in individuals with OAVS requires multidisciplinary surgical care throughout life. Its complex etiology remains poorly understood despite the many case reports and several associated genetic and environmental factors. In this study, we present the molecular findings of a family affected with macrostomia or abnormal mouth contour, preauricular tags, and uni- or bilateral uni- or bilateral ptosis. According to the original clinical description of this family, there are no hearing, spine, limbs, or renal defects ^1^.

The study of large families with incomplete penetrance and wide phenotypic variability allows us to search not only causal genes but also possible modifiers, both genetic and non-genetic. Although our results for this large family indicated that the disease follows an autosomal dominant inheritance; our analysis showed that missense mutations on the genes *SIX1*, *PDGFRA* and *KDR/VEGFR2* had the best evidence of segregation with the OAVS phenotypes in this family.

*SIX1* haploinsufficiency or hypomorphic mutations in *SIX* genes have been reported in patients with Branchio-oto-renal syndrome (BOR) (OMIM 113650), renal dysplasia, hearing loss, and frontonasal dysplasia syndrome^61^. BOR has craniofacial and vertebral signs similar to OAVS. Studies of *Six* genes in Drosophila show that the *Six* gene family is essential for eye morphogenesis ^62^; and in mice, *Six1* is required for normal development of the kidney, muscle and inner ear, midface and mandibular structures and is co-expressed with Eya factors ^33, 63–65^. *EYA* proteins interact with *SIX* proteins during development and the interaction causes *EYA* proteins to translocate to the nucleus and as a complex *SIX-EYA* can activate gene transcription^66^. Interestingly, *EYA1* is also involved in BOR and mutations in *EYA3* have been detected in patients with OAVS ^22, 23^. Moreover, a *DACH1* deletion and a *DACH2* duplication were recently described in two independent patients with OAVS^67^. These data support that dysregulation of the Retinal Determination Gene Network (*RDGN*) of genes also known in animals as the *PSEDN (PAX-SIX-EYA-DACH* network) which functions in eye development and others such as muscles, endocrine glands, placodes, and pharyngeal pouches is involved in the etiology of OAVS^68^.

In addition to the evidence from previous studies, our *in-silico* analysis for *SIX1* shows that the missense mutation D227E found in this family has the potential to interfere with the protein flexibility and its regulatory function. Lastly, Richieri-Costa pointed out that the external ophthalmoplegia presented by some family members that were affected with uni- or bilateral ptosis should be differentiated^1^. Notably, *Six 1* deficiency in mice causes reduced and disorganized muscle mass ^69^ and *Six 4* causes exacerbated craniofacial defects and severe muscle hypoplasia ^70^. Finally, a study with adult mice showed that *Six1* and *Eya1* are implicated in the establishment and control of the fast-twitch skeletal muscle phenotype^70^.

*PDGFRA* is Tyrosine-protein kinase that acts as a cell-surface receptor for platelet-derived growth factors *PDGFA, PDGFB* and *PDGFC* with a key role on regulation of embryonic craniofacial and neural crest development, palatogenesis and neural tube closure^71–73^. Mice homozygous but not heterozygous mutants for *Pdgfra* have cleft palate due to NCCS migration defects, however after ethanol exposure both homozygous and heterozygous *Pdgfra* mutans developed profound defects in the palate and pharyngeal skeleton related to neural crest apoptosis and suggesting a protective role against ethanol-induced craniofacial defects ^74^. Interestingly, a *PDGFRA* (c.C2740T; p.R914W) rare missense mutation was found to be functional with a dominant negative effect on a family with multiple affected with nonsyndromic OFC exhibiting decreased penetrance. This is in the same amino acid residue from our *PDGFRA* mutation R914Q. The wild-type (R914) forms direct hydrogen bonds with I909, Y913 and W933 plus three additional amino acids (I909, G912 and S935) ^75^ and it is likely that our mutation R914Q just like R914W obliterates some or all of these hydrogene bonds^76^. Lastly, in zebrafish, genetic alteration of *Pdgfra* caused unilateral clefts of the upper lip^76^, which could relate to the macrostomia defect which is also a form of facial cleft in the family presented.

The *KDR/VEGFR2* gene encodes for a Tyrosine-protein kinase that acts as a cell-surface receptor for *VEGFA, VEGFC* and *VEGFD*^77^ and mediates VEGF-induced endothelial proliferation, survival, migration, tubular morphogenesis and sprouting. The Vascular Endothelial Growth Factors (*VEGFs*) / receptors (*VEGFRs*) system plays an important role in angiogenesis, as well as osteogenesis, during bone development, growth, and remodeling, attracting endothelial cells and osteoclasts and stimulating osteoblast differentiation. *VEGFR2* mediates differentiation and proliferation of endothelial cells, and it is express in osteoblasts and osteoclasts, particularly during bone remodeling (Byun et al., 2007), Also *VEGFR1, VEGFR2* and VEGFR3 are expressed in both human and fetal and adult bone albeit with significantly higher expression in fetal samples especially in mandibles^78^. The higher expression of *KDR/VEGFR2* on fetal mandibles correlated well with the OAVS condition where microsomia and micrognathia are common occurrences. During mandibular development the neural crest cell-derived VEGF promotes adequate vessel growth and arterial stability and vessel-derived factors enable appropriate levels of chondrocyte proliferation and morphogenesis of Meckel’s cartilage, which are essential for mandibular enlargement^79^. The loss of VEGF reduces chondrocyte proliferation in Meckel’s cartilage and impairs jaw outgrowth. Cases with hemifacial microsomia also present malformations of the mandibular artery and reduction of mandibular medullary cavity volume that correlates with mandibular hypoplasia^79^. Finally, relevant to reconstructive surgery, *VEGFR2* is mostly produced during the early period of bone regeneration and modulation of *VEGF/VEGFR* could contribute to graft integration and improve outcomes for OAVS cases undergoing surgical reconstruction^80^.

Related to ocular phenotypes present in the OAVS condition, both *VEGF* and *KDR/VEGFR2* transcripts have correlated expression during the normal development of the ocular vasculature in humans during early development and in later stages *KDR* is linked to the development of the retinal vascular system^81^.

The presence of more than one likely pathogenic mutation in the same family raise different possibilities.(1) it confirms the complex etiology of the disease; (2) it is possible that one of the mutations is the main pathogenic mechanism and the additional mutations are modifiers or contributors to the phenotypic variability assuming the genes act on interacting pathways; (3) the multiple heterozygote mutations may be independent events, each one contributing to a distinct phenotype (“multiple hits”) yet their additive effects increases the risk for multiple phenotypes to simultaneously appear on an individual has seen in the craniofacial microsomia family with digenic inheritance due to mutations in *EYA3* and *EFTUD2*^82^. Complex, nontraditional models of inheritance should include incomplete penetrance, environmental and epigenetic factors.

Epigenomic studies can explain how genes and environmental exposures interact to affect craniofacial development and lead to disease phenotype.

The recent advances of whole genome technologies and the ability to integrate different OMICs platforms will likely shed light into the regulatory landscape underlying the OAVS, filling the gaps in genotype-phenotype correlations improving diagnosis patient care and paving the way for future prevention strategies.

## Data Availability

All data produced in the present work are contained in the manuscript

## Acknowledgments

Funding: Dewel Endowed funds University of Iowa Department of Orthodontics; NIDCR K01DE027995; Iowa Institute for Oral Health Research; College of Dentistry Student Research Program.

---

**Supplemental Table 1:**
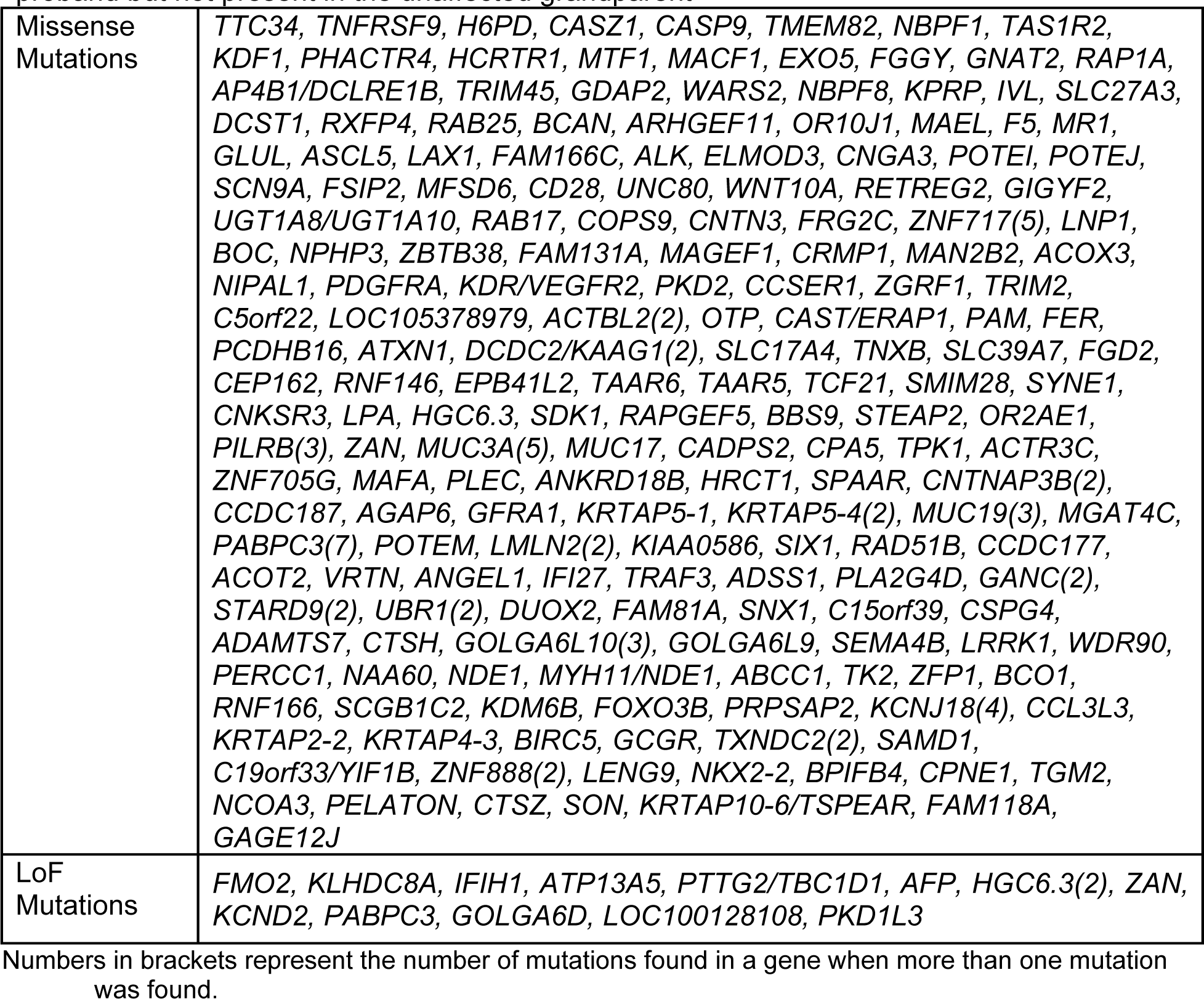
Genes with LOF and Missense mutations present in the parent and proband but not present in the unaffected grandparent.

## Footnotes

* The full pedigree is not included in this preprint to protect confidentiality of the patients. For additional information, please contact the corresponding author.

